# Are temperature suitability and socioeconomic factors reliable predictors of dengue transmission in Brazil?

**DOI:** 10.1101/2020.11.02.20224444

**Authors:** Lorena M. Simon, Thiago F. Rangel

## Abstract

**Background:** Dengue disease is an ongoing problem, especially in tropical countries. Like many other vector-borne diseases, the spread of dengue is driven by a myriad of climate and socioeconomic factors. Over recent years, mechanistic approaches have predicted areas of dengue risk according to the temperature effect on mosquitos’ lifespan and incubation period shaping their persistence and competence in transmission. Within developing countries such as Brazil, heterogeneities on socioeconomic factors are expected to create variable conditions for dengue transmission by its main vectors. However, both the relative role of socioeconomic aspects and its association with the temperature effect in determining the effective dengue prevalence are poorly understood.

**Methodology/Principal findings:** Here we gathered essential socioeconomic factors comprising demography, infrastructure, and urbanization over 5570 municipalities across Brazil and evaluated their relative effect on dengue prevalence jointly with a previously predicted temperature suitability for transmission. Using a simultaneous autoregressive approach (SAR), we showed that the variability in the prevalence of dengue cases across Brazil is highly explained by the combined effect of climate and socio-economic factors. Moreover, the temperature effect on transmission potential might be a better proxy at some dengue epidemy seasons but the socioeconomic factors are tightly linked with the recent increase of the dengue prevalence over Brazil.

**Conclusions/Significance:** In a large and heterogeneous country such as Brazil recognizing the drivers of transmission by mosquitoes is a fundamental issue to effectively predict and combat tropical neglected diseases as dengue. Ultimately, it indicates that not considering socioeconomic factors in disease transmission predictions might compromise efficient strategies of surveillance. Our study indicates that sanitation, urbanization, and GDP are regional indicators that should be considered along with temperature suitability for dengue transmission, setting a good starting point to effective vector-borne disease control.

**AUTHOR SUMMARY:** Dengue, a disease transmitted by mosquitoes, is a great problem in countries where the climate is predominantly hot and wet. Researchers know that temperature plays an important role in mosquitoes’ ability to transmits diseases. Usually, temperature alone is a good explanation for why dengue occurs in certain regions that have stable warm temperatures. Here we show that, in addition to the role of temperature on dengue spread, large urban areas with sanitation infrastructure and health assistance also prelude dengue cases prevalence. We highlight that dengue surveillance should consider socioeconomic regional differences. For instance, greater urban centers might be the focus of the dengue burden, where the presence of medical assistance and sanitation seems not to avoid the increase in disease cases. Conversely, less urbanized regions with suitable temperature for dengue transmission might require distinct actions for the disease combat.

## 1. INTRODUCTION

The presence and prevalence of many infectious diseases have clear geographic structures. These health threats vary from country-to-country and cause the loss of millions of lives annually (1,2). Identifying patterns and drivers of infectious diseases spread has become a fundamental concern on disease ecology (3). For instance, understanding why some regions have a higher charge in diseases and pathogens richness than others might help to identify hotspots for infections outbreak (2). However, a multitude of factors determines infectious disease geographical distribution and potential outbreaks, spanning from socioeconomic (*e.g*. urbanization; population density) to environmental (*e.g*. temperature; precipitation) and biotic (*e.g*. vectors competition) aspects (1,4). Acknowledging the variation over space in these drivers of infectious disease is increasing among ecologists in an attempt to identify regions of outbreak potential, once the disease dynamic is as tightly linked with exogenous factors as it is with endogenous mechanisms (5,6).

Dengue, a mosquito-borne infectious disease, is a global public health concern. The incidence of dengue has increased thirty-fold over the last five decades, and it is estimated that approximately one hundred million new infections occur annually (7). In the Americas, the disease is present in almost all countries with great prevalence (8), where rapid urban expansion led to ideal environmental conditions for dengue to spread (9). Nonetheless, dengue is still considered a neglected tropical disease (10). The geographic distribution of dengue vectors and the probability of virus transmission to human hosts are likewise driven by the ecological role of temperature (9,11). Despite its pervasiveness, the effect of ecological variables like temperature and precipitation might be outweighed by the influence of socio-economic aspects on dengue transmission (12), although ecological suitability approaches have mainly focused on the temperature effect on outbreaks potential (13,14).

On the one hand, climatic characteristics are important ecological drivers of most vector-borne infections (2,15), of which biological cycles and infectious stages are directly affected (16). For instance, the temperature can either reduce the infectious effectiveness by lowering the vector lifespan or increase infectiousness by shortening the incubation period (increasing epidemic chance) (5,15). In vector-borne diseases, the geographic range of pathogens and vectors is outlined by climate features due to its implication in traits such as reproduction, survival, and extrinsic incubation period, bounding their fitness (2,17,18). Accordingly, vectors’ (mostly arthropods) physiology and the exposure to pathogens are constrained by temperature, generally fitting in a thermal optimum (*i.e*., between the maximum and minimum tolerated) (19). The resulting efficiency in the vector-borne disease transmission is determined by the species responses to temperature bounds (6,11).

On the other hand, socio-economic factors are likewise critical in shaping the prevalence of infectious diseases (1,4,12). For instance, an increase in per capita healthcare expenditure is expected to decrease infectious disease prevalence (4), whereas the increase in human density relates to the augmentation of cases (20). Income and demography are frequently considered critical factors driving the spread of infectious diseases (1,4). A country’s income incorporates correlated factors that include sanitation, education, and health assistance, where the lack of investments is often associated with the rise of infectious disease prevalence (1,12). The population size, density, and urbanization are in turn demographic factors, of which increase raises the potential of transmission contact, also providing ideal microclimate and sites for vectors reproduction and development (16,20). In this sense, some infectious diseases prevail in countries where the characteristics of both demography and infrastructure create favoring conditions for transmission outcomes (12).

Distinct approaches have been used to address the presence and prevalence of infectious diseases, such as mechanistic (*i.e*., process-based) and statistical models (21,22). The mechanistic models rely on empirical information on disease transmission to estimate parameters in a bottom-up procedure (22). For instance, Brady *et al*. (23) used a mechanistic model to estimate the thermal limits of dengue through the relationship between temperature and *Aedes spp*. fitness. Most of the mechanistic approaches in mosquito-borne diseases have integrated the temperature effect on transmission traits to predict global geographical patterns, not considering lower scale socio-economic variations or considering it merely as additional factors (24). Recently, a multi-model climate-driven approach has also been proposed to forecast *Aedes*-borne diseases and support surveillance operations (25). Albeit integrating many transmission-related factors might turn intractable in a process-based procedure, the absence of key drivers still brings uncertainty to the estimated transmission potential (24).

Although dengue is present in almost all tropical and subtropical countries (7), Brazil has experienced a higher-than-expected number of infection cases in the last century (8,10). Since the 80’s, the reintroduction of dengue in the country has led to its rampant geographic expansion (26). Initially the presence of dengue virus was more intense in large urban centers, but since 90’s it has spread to small towns and countryside. In the 2000s, the dengue vectors (*i.e*., *Aedes aegypti* and *Aedes albopictus*) were already present in 72% of Brazilian municipalities, dramatically increasing the disease cases and overloading the Brazilian health system (27). Although pervasive across Brazil, the prevalence of dengue cases varies widely among the urban centers making it difficult for dengue season preparedness especially in conjunction with other infectious disease outbreak (27,28).

In this paper we evaluate the relative impact of socioeconomic conditions and temperature suitability over the spatial pattern of dengue fever prevalence across Brazil. We used a previously estimated temperature suitability for dengue transmission (23) and 7 socio-economic variables to study the prevalence of dengue disease in 5570 municipalities across Brazil. We also seek to understand the fit between the estimated temperature suitability for transmission and the effective prevalence of dengue in the last years. We predict that in a highly heterogeneous country, such as Brazil, socioeconomic factors are the greater source of high levels of dengue prevalence. In Brazil, regions with the highest dengue prevalence are not those with highest estimated temperature suitability for transmission, although suitability is still a good indicator of the disease occurrence.

## 2. MATERIALS AND METHODS

### (a) Data

#### (i) Dengue cases

The number of dengue cases in Brazilian municipalities from 2007 to 2013 were obtained from DATASUS, a national public health database maintained by the Brazilian Health System (29). From 2014 to 2016 we obtained, from the same source, the number of probable cases (as opposed to diagnosed cases), which has a higher sensitivity for reports during epidemic periods. The dengue cases are notified as probable based on clinical and epidemiological evidence, and are carried out by a local health surveillance team (30).

### (ii) Temperature suitability

Here we used the simulated dengue transmission suitability maps by Brady *et al*. (23) as a predictor of dengue presence and prevalence in Brazil, from which we extracted the raster information regarding each municipality. Brady *et al*. (23), estimated the dengue transmission suitability from the temperature influence on mosquitoes’ (*Ae. aegypti* and *Ae. albopictus*) survivorship and extrinsic incubation period (*i.e*., EIP). The EIP represents the period between mosquito biting an infected host and being able to transmit the virus after processing the pathogen into the gut (*i.e*., become infectious) (31). Brady’s *et al*. (23) mechanistic model considered the dynamic between the EIP and the adult vector survival - both temperature-dependent - over the basic reproductive number (*i.e*., R_0_) (see (32,33)). The model outcome was then combined with a spatially explicit temperature data, producing predictive maps of vectors’ suitability range on the persistence and competence of dengue transmission (23).

#### (iii) Socio-economic drivers

For each Brazilian municipality, we likewise gathered important socio-economic predictors to the distribution and prevalence of mosquito-borne diseases (see Table A in S1 Appendix), which were: human population density, urbanization, population size, amount of health facilities, gross domestic product (henceforth GDP), education and sanitation. The referred socio-economic variables and the political-administrative division map of Brazilian municipalities were obtained from IBGE (34,35). The Brazilian political-administrative extension comprises 5570 municipalities, which were all included.

Epidemiological studies suggest that regions with high population density and great urbanization favor the increase of dengue cases by facilitating vectors’ mobility and reproduction (20,36). Here we estimated the population density as the ratio among population size and the area of each municipality. We accounted for the population size as the total number of people within each municipality, following the IBGE classification. To access the proportion of urbanization within municipalities, we estimated the ratio between the urbanized area (maps based on satellite images (34)) and the whole political-administrative extent from each municipality. Also, to account for the effect of medical diagnosis, notification and local investments, we used the number of people assisted by educational and health assistance in each municipality (16,27). Finally, to represent economic development, we also considered GDP (log scale) and the presence of the basic sanitation system (*i.e*., sewage treatment and/or rainfall water management) (see Table A in S1 Appendix).

### (b) Analyses

We employed linear correlations among the predictor variables to assess their collinearity. In a stepwise procedure, we evaluated the non-independence between the predictors by measuring the Variance Inflation Factor (VIF) among them and set apart the most redundant variables. We started with a full model and iterated the procedure until all the predictors had a VIF lower than 10 (37). According to this procedure the variables population size and education are the most collinear and were therefore removed from analyses. Although important to disease transmission in general (Table A in S1 Appendix), the population size has a confounding relation with GDP in Brazil as a consequence of internal migration patterns to economically developed areas (38).

Because our analysis is spatially explicit, we accounted for the spatial autocorrelation that could potentially inflate Type-I errors of statistical inferences (39). We used a simultaneous autoregressive model (SAR), which comprises linear regressions with the addition of an autoregressive term specifying the strength of dependence between each pair of locations. We followed the Kissling & Carl (40) recommendation of reliability and better performance of SAR_error_ model, and also used the row standardized coding for the spatial weight matrix, all using the R packages spdep (41) and spatialreg (42). Finally, we examined Moran’s I correlogram from models’ residuals to ensure the effective control of the spatial autocorrelation.

To test for the dissimilarity between putative drives of dengue prevalence, once the period of 2015-2016 had higher disease prevalence than previous years (8), we implemented two separate SAR_error_ models using the log of dengue cases for the periods of 2007-2014 and 2015-1016.

Because our SAR_error_ models that are estimated by maximum likelihood the calculation of coefficient of determination is different from standard OLS regression (43). We therefore estimated coefficients of determination (R^2^) through the following formula:

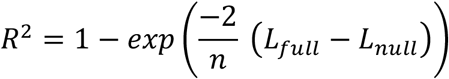

where, *n* = sample size, L_*full*_= likelihood of the fitted model, L_*null*_= likelihood of the null model-the model containing no autoregressive term -. All analyses and maps were performed using R 3.5.0 (44) (https://www.r-project.org).

## 3. RESULTS

Dengue cases are unevenly distributed across Brazil, both in occurrence and prevalence (*i.e*., number of cases) (Fig 1). Over the last years, most dengue cases showed to be concentrated into the Southeast and Midwest regions of Brazil, but were also less frequently present in the North and Northeast. From 2007-2014 (Fig 1a) there were fewer recorded dengue cases when compared with a more recent epidemic period (2015-2016; Fig 1b), albeit reaching the Northern region with a higher prevalence. From 2015 to 2016, the dengue prevalence was greater in the Southeast, Midwest, and Northeast regions of Brazil, where the total number of cases almost doubled proportionally to the previous seven years on which dengue was predominant into the Southeast.

**Fig 1.**
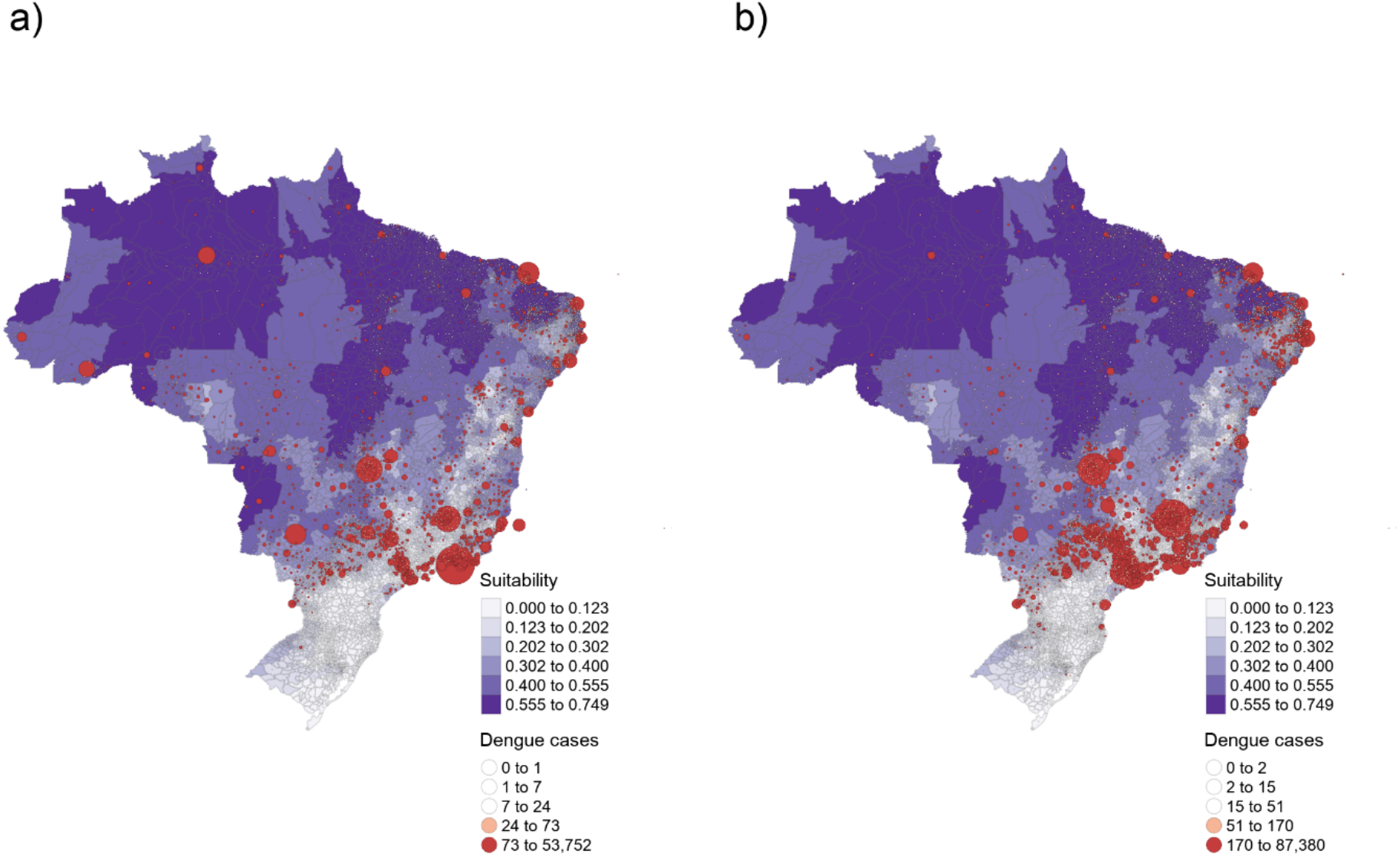
Brazilian dengue cases distribution of (a) 2007 to 2014 and (b) 2015 to 2016. The size of the red circles corresponds to the magnitude of dengue prevalence (*i.e*., the mean number of cases). The intensity of purple indicates the mean temperature suitability for the dengue transmission by *Aedes* spp. vectors (23).

The graphical comparison between the distribution of real dengue cases (Fig. 1, red circles) and the estimated temperature suitability for potential dengue transmission (Fig. 1, purple shades) showed a lack of correspondence in both periods. Although the Brady’s *et al*. model predicts high temperature suitability for dengue transmission in the North and Northeast regions of Brazil, fewer dengue cases truly occurred within this extensive area. Conversely, most dengue cases were recorded into the Southeast and Midwest regions, where the model estimated lower temperature suitability. Notably, the greatest proportion of dengue cases are concentrated in areas where the model did not predict temperature suitability for dengue transmission. However, the temperature suitability model accurately predicts the decreased potential for dengue outbreak in southern Brazil, where the number of autochthonous dengue cases was lower in the whole period of almost ten years.

The autoregressive model revealed that the relative importance of the socio-economic factors and estimated temperature suitability for dengue transmission in Brazil (Table 1). From 2007 to 2014, the urbanization (as well as its association with the temperature suitability), health facilities and GDP were the socioeconomic factors that best explained the number of dengue cases across Brazil. In contrast, GDP and sanitation were the solely socioeconomic factors that accounted for the disease distribution and prevalence in 2015-2016. However, in both 2007-2014 (z = 67.423) and 2015-2016 (z = 58.691), GDP was the predictor that best explained number recorded dengue cases across the country. Albeit in less magnitude, temperature suitability for dengue transmission also showed higher explanatory power for the distribution of dengue cases in 2007-2014 (z = 14.825) than in 2015-2016 (z = 7.145).

**Table 1.** SAR_error_ results for both periods, previous (2007 to 2014) and recent (2015 to 2016).

| Dependent Variable: Previous dengue cases (ln) |  |  | Dependent Variable: Recent Epidemic dengue cases (ln) |  |  |
| --- | --- | --- | --- | --- | --- |
| Independent Variables | Parameter Estimate (Standard Error) | z Value | Independent Variables | Parameter Estimate (Standard Error) | z Value |
| Temperature Suitability | 3.858 (0.26)*** | 14.82 | Temperature Suitability | 2.145 (0.30)*** | 7.14 |
| Urbanization | -0.108 (0.27)*** | -2.34 | Urbanization | 0.061 (0.31) | 0.19 |
| Population Density | 1.016e-04 (4.97e-05)* | 2.04 | Population Density | 7.875e-05 (5.81e-05)* | 1.35 |
| Health Facilities | 1.483e-04 (3.25e-05)*** | 4.56 | Health Facilities | 7.824e-05 (3.81e-05)* | 2.05 |
| GDP (ln) | 0.796 (0.01)*** | 67.42 | GDP (ln) | 0.811 (0.01)*** | 58.69 |
| Sanitation | 0.047 (0.03)* | 1.56 | Sanitation | 0.142 (0.03)*** | 4.05 |
| Temperature Suitability vs. Density | -1.115e-04 (1.829e-04) | -0.61 | Temperature Suitability vs. Density | -1.766e-04 (2.14e-04) | -0.83 |
| Temperature Suitability vs. Urbanization | 4.448 (1.12)*** | 3.98 | Temperature Suitability vs. Urbanization | 1.723 (1.30)* | 1.32 |
| R<sup>2</sup> | 0.53 |  | R<sup>2</sup> | 0.49 |  |
| AIC (lm) | 19917 |  | AIC (lm) | 21264 |  |
| AIC (SAR<sub>error</sub>) | 15744 |  | AIC (SAR<sub>error</sub>) | 17475 |  |
| Moran's I (p-value) | (0.99) |  | Moran's I (p-value) | (0.99) |  |

\* $p \leq 0.10$ .
\*\*\* $p \leq 0.01$ .
(ln) Natural log.

Human population density was not a significant explanatory factor for the number of recorded dengue cases in both periods, neither was its interaction with estimated temperature suitability for dengue transmission. However, urbanization, a proxy for the expansion of human-modified areas, was found to be an important predictor of the dengue cases from 2007 to 2014 (Table 1). Both urbanization itself and its interaction with the temperature suitability for dengue transmission were significant in those previous years, but less relevant in the more recent period of 2015-2016. The coefficients of determination (R^2^) of the SAR_error_ models varied between periods, suggesting that the same socio-economic variables and the temperature suitability for dengue transmission have higher explanatory power during years of lower transmission (R^2^ = 0.53) than in periods of an increased outbreak (R^2^ = 0.49). The SAR model had lower AICs than its Ordinary Least Squares (OLS) counterpart (Table 1).

Spatial autocorrelation was successfully controlled by the SAR model (Fig A in S2 Appendix). The residuals of the models revealed a narrow variation on the range of values and there were no marked spatial patterns of residuals across Brazil (Fig 2). In the southern region the residuals are very small in many municipalities, indicating that model predictions were accurate in these areas. In contrast, some municipalities in the Amazon region had negative residuals (overestimated number of dengue cases), whereas some municipalities in central and southeast Brazil had positive residuals (underestimated number of dengue cases) (Fig 2).

**Fig 2:**
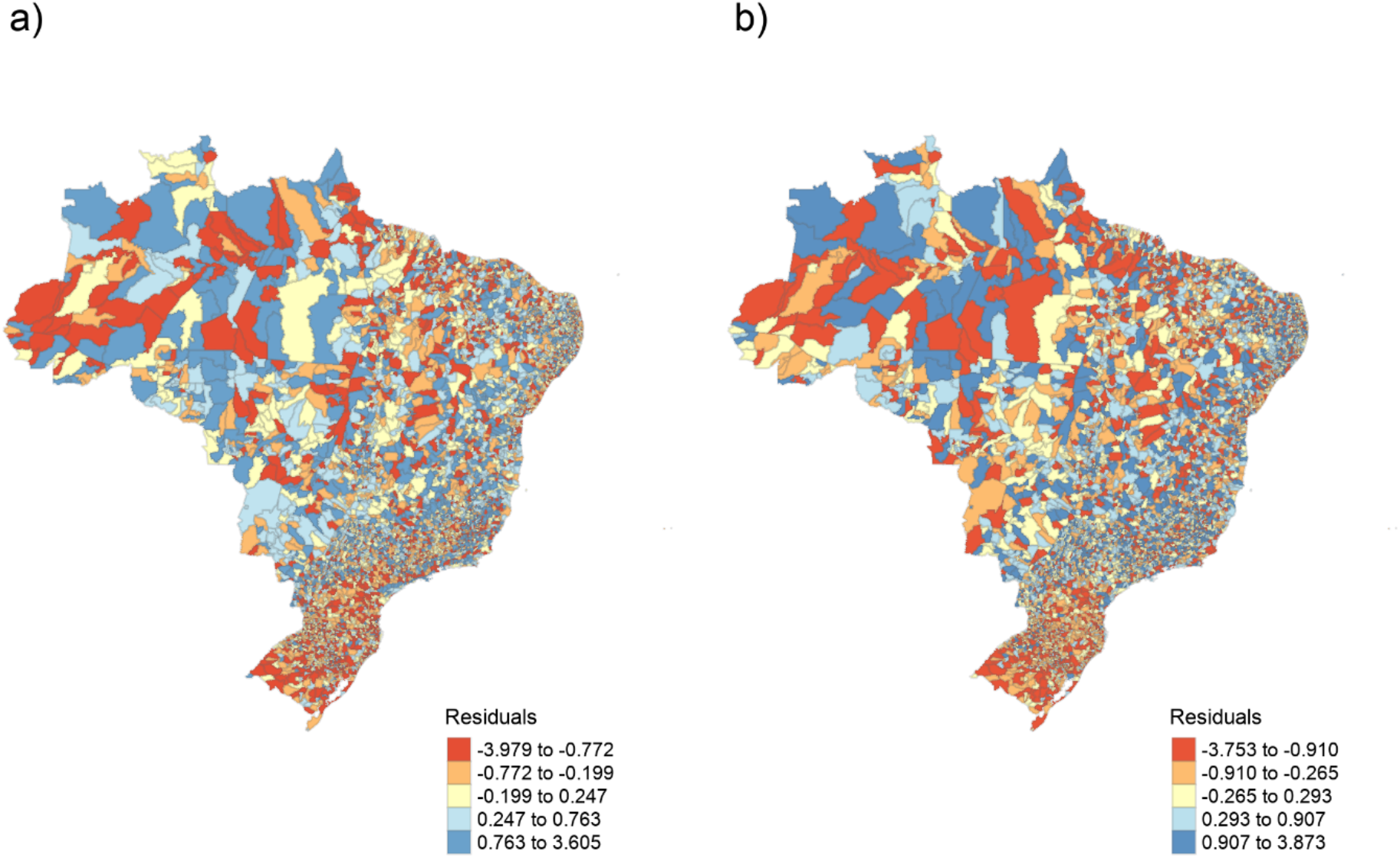
SAR_error_ models’ resulting residuals. (a) 2007 to 2014 and (b) 2015 to 2016, spatially distributed by municipalities.

## 4. DISCUSSION

The higher prevalence of dengue over Brazil compared to other countries has intrigued researchers for decades, revealing that singular factors might regulate the transmission of this infectious disease within particular countries (27). This constant reemergence and maintenance of a high number of dengue cases in Brazil remain unclear, which is justified due to the complex nature of the biological features of virus (*e.g*., serotypes, virulence), host (*e.g*., immune system) and vectors (*e.g*., ambient suitability, reproduction rates) (45). Therefore, the magnitude of the dengue incidence fluctuates according to a myriad of environmental factors. Our results showed that, although temperature suitability for transmission is a good indicator of dengue occurrence, the socio-economic characteristics are the fundamental determinants of the spatial patterns in dengue prevalence in Brazil.

Similar to other tropical nations, Brazil is a heterogeneous country that has undergone substantial urban growth in recent decades. This rapid urban expansion along with favorable climatic conditions creates an ideal scenario for the establishment and spread of critical infectious diseases, especially those carried by mosquitoes (22). However, dengue occurrence and number of cases differ substantially among regions within Brazil. Thus, predicting the potential transmission of mosquito-borne infectious disease requires incorporating environmental and socio-economic heterogeneities among regions, especially in countries where an endemic scenario is well established (27). Although the temperature is a physical factor known to affect the physiology of mosquitoes and its capacity as disease vectors (18), here we demonstrated that the predicted temperature suitability for dengue transmission does not account for the largest proportion of variation in the number of dengue cases across Brazil, where socioeconomic factors showed fundamental in promoting or preventing dengue transmission.

Because temperature drives the suitability for dengue transmission (25) global climate change may substantially alter the spatial pattern in the distribution and prevalence of dengue (11,46). In countries where the autochthonous transmission of dengue is established, the rising temperatures may intensify transmission by favoring vectors’ survival, reproduction, and biting rates (6). Estimates of dengue transmission suitability under temperature trends have been made as an attempt to anticipate epidemics spread and to plan mitigation strategies (22,23,47). Frequently, such estimates point to a potential increase of the dengue burden under current and future temperature scenarios. Under lower spatial and temporal scales, the relationship between temperature and multiple other drivers, such as urbanization, GDP, and sanitation, should be more appropriate to describe patterns of disease transmission (16,18). For instance, we showed that Brady’s *et al*. model point to a high dengue transmission potential in the north of Brazil (*e.g*. Amazon region), although few cases were reported there; this reveals that, under the temperature perspective, their model may correctly predict the potential of dengue transmission in this area, but the effective transmission of dengue depends on socio-economic factors, such as demography. Notwithstanding, the thermal optima for dengue transmission are limited considering the unimodal effect of temperature (6,11). Increasing the temperature over the tolerance of vectors and pathogens might instead decrease dengue transmission in response to microclimates modulation on urban sites (28), justifying an overestimated number of dengue cases in some cities.

After controlling for the role of temperature suitability for transmission, our findings highlight that the socio-economic conditions contribute substantially to dengue prevalence. Jointly, the GDP and urbanization surpassed the importance of temperature suitability during previous years, whereas the GDP along with sanitation were determinants of the recent dengue increase. Although temperature constrains the fitness of dengue vectors, environmental conditions created by urban settings still modulate their persistence and competence (48). Conversely, larger urban centers are usually equipped with more health facilities, increasing the capacity for dengue diagnosis and reports. Higher economic income along with expected development might also reduce infectious diseases by increasing the sanitation system and vector combat (12,49). However, our results pointed out a positive relationship between the sanitation system and GDP with the prevalence of dengue, indicating that their presence by itself may not attest to the benefits of economic wealth in reducing the dengue disease cases in Brazil.

The demography in urban environments is thought to be an important driver of dengue prevalence (50). For instance, a temporal analysis of the dengue outbreak in Singapore found that the population demography is the main driver for the dengue increase in the last years (51). This encounter is usually accurate given the expected mixed contact between hosts and vectors, where the pure increase in individuals density is expected to increase the contact rates between hosts and vectors (32). However, after controlling for other covariates, our model did not find a substantial effect of demography on the prevalence of dengue cases across Brazil. Although demography might be a good proxy for the number of susceptible individuals, in dengue-endemic countries such as Brazil the serotypes immunization is likely to reduce this proportion (52). Still, the population density may have great importance at the local scale (*e.g*., among neighborhoods), once it increases the probability of vector contact with hosts when the virus is installed (53). The greater relation of municipalities’ GDP with dengue prevalence could also be an indication of the interchange between higher population, immunization, and different serotypes circulating, once Brazilian cities with greater income grown faster by historically being attractive for migration (54).

There is no doubt that the burden of dengue is heavier in some regions than others. In Brazil, where dengue cases greatly vary across space and time, we highlight that the combined effect of climate and socio-economic factors drive the dengue occurrence and prevalence. Still, due to the lack of reliable reports on dengue prevalence, most predictive models overemphasize the role of temperature on dengue transmission in large scales (22). By accounting for the effect of socio-economic drivers in an extremely heterogeneous country, we showed that dengue prevalence is explained not only by the temperature suitability for transmission but also by social and economic factors. Highly urbanized centers with great income were found to be epicenters of dengue transmission in Brazil, aligned with other infectious diseases (55). Consequently, projections of dengue risk areas over actual or future climatic conditions should include socioeconomic covariates to make predictions reliable for decision making regarding a vector-borne tropical neglected disease such as dengue, especially when considering that dengue season might come when another infectious disease epidemic is already overloading the health system (*e.g*. SARS-CoV-2). Here we emphasize the need of considering social, economic, and cultural differences between Brazilian regions for effective decision making.

## Data accessibility

This article has no additional data.

## Data Availability

All relevant data are within the manuscript and its Supporting Information files.

## Acknowledgments

This study was financed in part by the Coordenação de Aperfeiçoamento de Pessoal de Nível Superior - Brasil (CAPES) - Finance Code 001.

## Supporting information

**S1 Appendix. Table of description and predictions to each socioeconomic variable used as an independent variable**.

**S2 Appendix. Figure showing the Moran’s I correlogram for** SAR_error_ **model’s residuals**

