## Supplementary material for "Are temperature suitability and socioeconomic factors reliable predictors of dengue transmission in Brazil?": S1 Appendix

### Supporting information: S1 Appendix

#### Are temperature suitability forecast and socioeconomic features reliable predictors of dengue transmission over Brazil?

Lorena M. Simon, Thiago F. Rangel

**This PDF file includes:**

Table A
References

**Table A. Table of description and predictions to each socioeconomic variable used.**

| Variables | Description | Prediction |
|---|---|---|
| Urbanization | The proportion of urbanized sites about the total municipality extent | Provides ecological conditions and reproduction sites, favoring vector proliferation and its closeness with population (Kuno, 1995; Gubler, 2011) |
| Population Density | Ratio between population size and municipal area | Flight capacity of Aedes mosquitos is limited, thus favored in densely human population (Kuno, 1995). |
| Population size | Total number of people residing in each municipality | The minimum population size required to sustain dengue transmission is over 10,000 (Kyle & Harris, 2008). |
| Education | People with access to school | Indicates the access to surveillance and prevention information, fundamental to strategies against dengue transmission (Teixeira *et al*., 2009). |
| Health assistance | Number of health establishments available | Greater access to medical care increases disease cases notifications and reports (Kuno, 1995). |
| GDP (gross domestic product) | Economic activity of each locality | Spending on the health system and in avoidance of disease vectors is closely related to local income (Bonds, 2012). |
| Sanitation system | Presence or absence of sewerage system and/or pluvial water system | The sanitation system absence provides vectors breeding sites (Kuno, 1995). |
