## Supplementary material for "Are temperature suitability and socioeconomic factors reliable predictors of dengue transmission in Brazil?": S2 Appendix

### Supporting information: S2 Appendix

Are temperature suitability forecast and socioeconomic features reliable predictors of dengue transmission over Brazil?

Lorena M. Simon, Thiago F. Rangel

**This PDF file includes:**

Fig. A

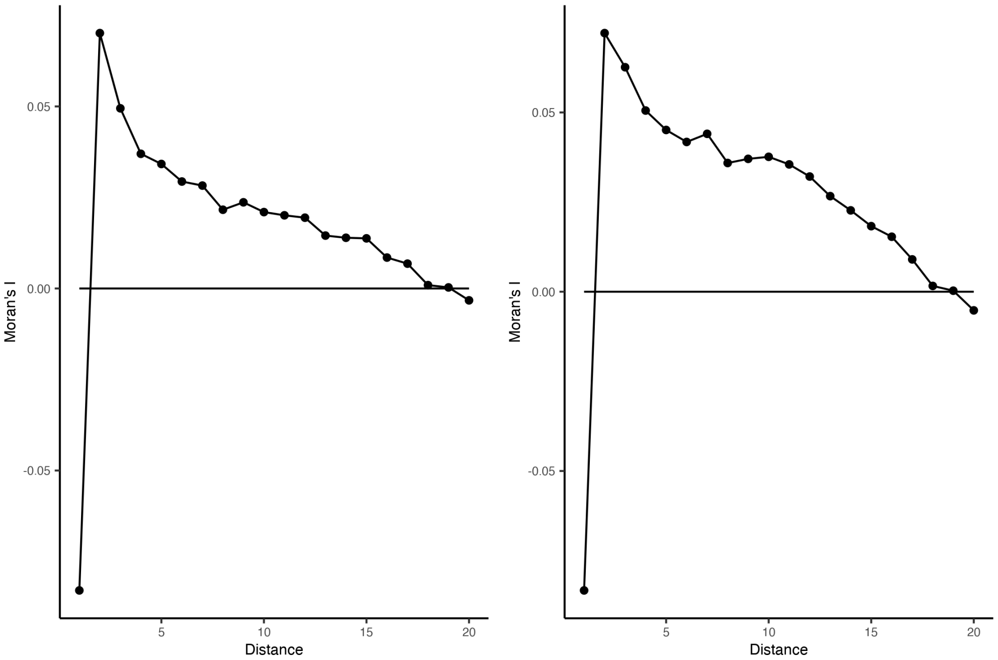


**Fig. A**

The Moran's I correlogram of $SAR_{error}$ model's residuals for 2007-2014 (Left) and 2015-2016 (right) periods of dengue. After accounting for the spatial autocorrelation the Moran's I value drops to 0 around 17 distance classes. To build the distance matrix we used the adjacency criterium between municipalities.
